# Genome-wide association study reveals loci with sex-specific effects on plasma bile acids

**DOI:** 10.1101/2022.12.16.22283452

**Authors:** Arianna Landini, Dariush Ghasemi-Semeskandeh, Åsa Johansson, Shahzad Ahmad, Gerhard Liebisch, Carsten Gnewuch, Regeneron Genetics Center, Gannie Tzoneva, Alan R. Shuldiner, Andrew A. Hicks, Peter Pramstaller, Cristian Pattaro, Harry Campbell, Ozren Polašek, Nicola Pirastu, Caroline Hayward, Mohsen Ghanbari, Ulf Gyllensten, Christian Fuchsberger, James F. Wilson, Lucija Klarić

## Abstract

Bile acids are essential for food digestion and nutrient absorption, but also act as signalling molecules involved in hepatobiliary diseases, gastrointestinal disorders and carcinogenesis. While many studies have focused on the genetic determinants of blood metabolites, research focusing specifically on genetic regulation of bile acids in the general population is currently lacking. Here we investigate the genetic architecture of primary and secondary bile acids in blood plasma, reporting associations with both common and rare variants. By performing genome-wide association analysis (GWAS) of plasma blood levels of 18 bile acids (N = 4923) we identify two significantly associated loci, a common variant mapping to *SLCO1B1* (encoding a liver bilirubin and drug transporter) and a rare variant in *PRKG1* (encoding soluble cyclic GMP-dependent protein kinase). For these loci, in the sex-stratified GWAS (N♂ = 820, N♀ = 1088), we observe sex-specific effects (*SLCO1B1* β ♂ = -0.51, *P* = 2.30×10^−13^, β♀ = -0.3, *P* = 9.90×10^−07^; *PRKG1* β ♂ = -0.18, *P* = 1.80×10^−01^, β ♀ = -0.79, *P* = 8.30×10^−11^), corroborating the contribution of sex to bile acid variability. Using gene-based aggregate tests and whole exome sequencing, we identify rare pLoF and missense variants potentially associated with bile acid levels in 3 genes (*OR1G1, SART1* and *SORCS2*), some of which have been linked with liver diseases.

## Introduction

Bile acids (BAs) are synthesised from cholesterol in the liver and subsequently stored in the gallbladder. After ingestion of food, BAs are secreted into the small intestine, where they contribute to the digestion of lipid-soluble nutrients^1^. Approximately 95% of BAs are then re-absorbed by the intestinal epithelium and transported back to the liver via the portal vein - a process termed “enterohepatic circulation”^2^. Primary bile acids in humans consists of cholic acid (CA), chenodeoxycholic acid (CDCA), and their taurine- or glycine-bound derivatives (TCA and TCDCA, GCA and GCDCA). Once secreted in the lower gastrointestinal tract, primary BAs are heavily modified by the gut microbiota to produce a broad range of secondary BAs, with deoxycholic acid (DCA), a CA derivative, and lithocholic acid (LCA), a CDCA derivative, being the most prevalent^2^. Bile acids also act as hormone-like signalling molecules, serving as ligands to nuclear (hormone) receptors. Through activation of these diverse signalling pathways, BAs control not only their own transport and metabolism, but also lipid and glucose metabolism, and innate and adaptive immunity^3^. Bile acids are thus involved in regulating several physiological systems, such as fat digestion, cholesterol metabolism, vitamin absorption, and liver function^4^. In addition, given their role in coordinating bile homeostasis, biliary physiology and gastrointestinal functions, impaired signalling of BAs is associated with development of hepatobiliary diseases, such as cholestatic liver disorders, cholesterol gallstone disease and other gallbladder-related conditions^5^, and of inflammatory bowel disease^6^. Further, bile acids have been implicated in carcinogenesis - specifically oesophageal, gastric, hepatocellular, pancreatic, colorectal, breast, prostate and ovarian cancer - both as pro-carcinogenic agents and tumour suppressors^7^. Thanks to their role as signalling molecules, BAs have been considered as possible targets for the treatment of metabolic syndrome and various metabolic diseases^8^. Further, BAs are able to facilitate and promote drug permeation through biological membranes, making them of general interest for drug formulation and delivery^9^.

While many studies have focused on the genetic determinants of blood metabolites^10–15^, research focusing specifically on bile acids in a large sample from the general population is currently lacking. Here we investigate the genetic architecture of primary and secondary BAs, reporting associations with both common and low-frequency/rare variants. First, we performed a genome-wide association meta-analysis (GWAMA) of plasma blood levels of 18 BA traits (N=4923). For a subset of this sample (female N=1088, male N=820), we perform sex-stratified GWAMA, to describe sex-specific genetic contributions to BA variability. We then explore whether complex traits or diseases have a role in influencing BA variability by using Mendelian Randomisation. We finally employ multiple gene-based aggregation tests to investigate rare (MAF < 5%) predicted loss of function (pLoF) and missense variants from whole exome sequencing affecting the 18 BA traits in a subset of our cohorts (N=1006).

## Results

### Loci associated with serum levels of bile acids

To investigate the genetic control of bile acids, we performed a GWAS meta-analysis on five cohorts of European descent (N = 4923), studying the associations of blood plasma levels of 18 primary and secondary bile acid traits with HRC-imputed genotypes/whole exome sequence data. Based on the number of below limit-of-detection (LOD) measurements, BAs were analysed either as quantitative or binary traits (Supplementary Table 1). In addition, two analysis approaches were carried out in parallel for quantitative traits: in one case, <LOD values were considered as missing, in the other case, they were imputed (Methods). An additive linear model was assumed for each bile acid trait, followed by fixed-effect inverse-variance meta-analysis. Overall, we identified 2 loci that passed the significance threshold (p-value < 3.57 × 10^−9^, Bonferroni adjusted for the number of independent bile acid traits) (Figure1), near the *SLCO1B1* and *PRKG1* genes. The most strongly associated locus (p=1.14 × 10^−16^), on chromosome 12 near *SLCO1B1*, showed consistent directionality across 4 of the 5 populations (Table 1), with the effect allele T of the sentinel SNP, rs4149056, associated with decreased serum levels of GDCA (quantitative). In the same locus, we found GLCA and the imputed GDCA trait to be significantly associated with the rs73079476 variant (Supplementary Table 2), in high linkage disequilibrium with the sentinel SNP, rs4149056 (r^2^ = 0.97). On the other hand, rs146800892, the sentinel SNP on chromosome 10 near *PRKG1*, has a minor allele frequency (MAF) lower than 1% in any cohort but CROATIA-Vis and might thus represent a cohort-specific association with GCA (Supplementary Table 2).

**Table 1:** Loci genome-wide significantly associated with at least one of the 18 bile acid traits in all samples and sex-stratified GWAMA.

| **All samples** |  |  |  |  |  |  |  |  |  |  |
| --- | --- | --- | --- | --- | --- | --- | --- | --- | --- | --- |
| **Locus** | **Gene** | **SNP** | **EA** | **OA** | **EAF** | **Beta** | **P** | **SE** | **N** | **Lead BA** |
| 12:20994540-21463812 | *SLCO1B1* | rs4149056 | T | C | 0.839 | -0.25 | 1.10x10<sup>-16</sup> | 0.03 | 4547 | GDCA |
| 10:53832549-53832549 | *PRKG1* | rs146800892 | T | C | 0.988 | -0.96 | 3.30x10<sup>-09</sup> | 0.16 | 900 | GCA |
| **Sex-stratified** |  |  |  |  |  |  |  |  |  |  |
| **Locus** | **Gene** | **SNP** | **EA** | **OA** | **Beta M** | **Beta F** | **P M** | **P F** | **N (M/F)** | **Lead BA** |
| 12:20994540-21463812 | *SLCO1B1* | rs73079476 | A | C | -0.51 | -0.31 | 2.30x10<sup>-13</sup> | 9.90x10<sup>-07</sup> | 820/1088 | GDCA |
| 10:53832549-53832549 | *PRKG1* | rs117834398 | T | G | -0.18 | -0.79 | 1.80x10<sup>-01</sup> | 8.30x10<sup>-11</sup> | 820/1088 | GCA |
| Each locus is represented by the SNP with the strongest association in the region, according to the p-value rejecting the null hypothesis of no association with at least one of 18 bile acid traits. In all samples analysis, an association was considered significant if the p-value was lower than $3.57 \times 10^{-9}$, the genome-wide significance threshold Bonferroni-corrected for the number of independent bile acid traits. In sex-stratified analysis, an association was considered significant if the p-value was lower than $5 \times 10^{-9}$, the genome-wide significance threshold Bonferroni-corrected for the number of independent bile acid quantitative traits. The two SNPs in the *SLCO1B1* locus are in high LD (LD $r^2 = 0.97$), while the two SNPs in the *PRKG1* locus represent two distinct signals (LD $r^2 < 0.001$).<br>Locus - coded as 'chromosome: locus start–locus end' (GRCh37 human genome build); Gene - suggested candidate gene; SNP - variant with the strongest association in the locus; EA - SNP allele for which the effect estimate is reported; OA - other allele; EAF - frequency of the effect allele; Beta - effect estimate for the SNP and bile acid with the strongest association in the locus; SE - standard error of the effect estimate, P - p-value of the effect estimate (two-sided Wald test with one degree of freedom); N - sample size; Lead BA - bile acid with the strongest association to the reported SNP; M - male specific analysis; F - female specific analysis. |  |  |  |  |  |  |  |  |  |  |

### Sex-specific associations of bile acid serum levels

To investigate whether the genetic component influencing bile acid variation may differ between men and women, we performed sex-specific GWAS meta-analysis of the 14 quantitative (imputed) bile acid traits for ORCADES and CROATIA-Vis cohorts (female N=1088, male N=820) and discovered two sex-specific associations. The association of GDCA with rs73079476 from the *SLCO1B1* locus was significant in male-only GWAS (beta = -0.51, p-value = 2.28 × 10^−13^) (Table 1, Figure 2A). The signal for the same locus in female-only GWAS, while consistent in terms of directionality, has a smaller effect size than in male-only analysis (beta = -0.31) and does not reach the significance threshold (p-value = 9.86 × 10^−7^), despite the slightly higher sample size (Figure 2). This suggests that the genetic effect of *SLCO1B1* locus on the serum levels of GDCA is larger in men than women. We also identified a sex-specific association of GCA at the *PRKG1* locus. In contrast to *SLCO1B1*, the sentinel SNP in *PRKG1*, rs117834398, has a larger effect in females than in males (female beta = -0.79, male beta = -0.18), and passed the significant threshold only in the female-specific analysis (female p-value = 8.26×10^−11^, male p-value = 1.81×10^−1^) (Table 1, Figure 2B). Interestingly, the sentinel SNPs at the *PRKG1* locus for the full meta-analysis and for the female-specific analysis are in linkage equilibrium (r^2^<0.01) and represent two independent associations in that locus. Overall, none of the significant association identified in one sex was replicated in the other, suggesting that the genetic contribution to serum BA levels is likely to be different in males and females. We have identified 13 additional associations (p-value < 5 × 10^−9^, Bonferroni adjusted for the number of independent quantitative bile acid traits) that might have sex-specific effects (Supplementary Table 3, Supplementary Figure 1). However, given the low allele frequencies and allele counts in the two analysed cohorts, further analyses are required to replicate these associations.

**Figure 1.**
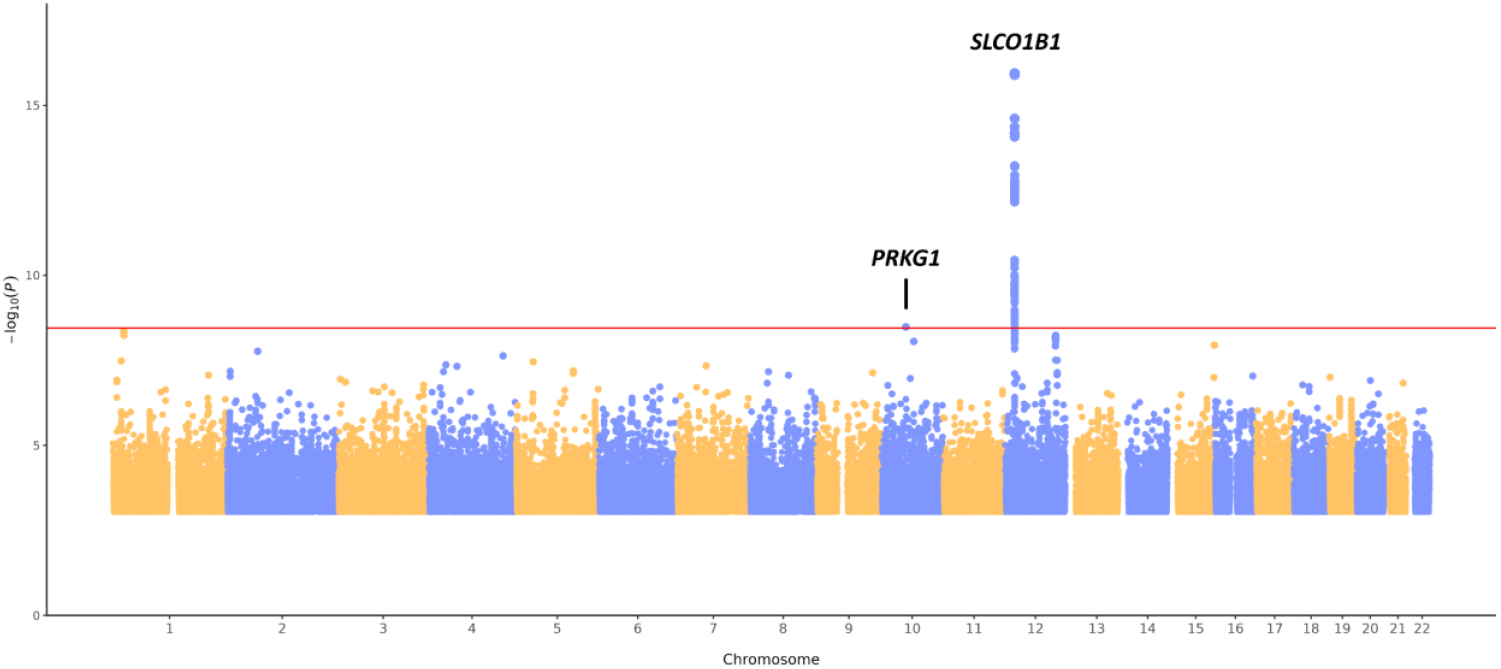
Summary Manhattan plot pooling together meta-analysis results obtained across 18 bile acid traits. The pooling was performed by selecting the lowest *p* value (y-axis) from the 18 bile acids for every genomic position (x-axis). The Bonferroni-corrected genome-wide significance threshold (horizontal red line) corresponds to 3.57 × 10^−9^. For simplicity, SNPs with *p* value > 1 × 10^−3^ are not plotted. *P* values are derived from the two-sided Wald test with one degree of freedom.

**Figure 2.**
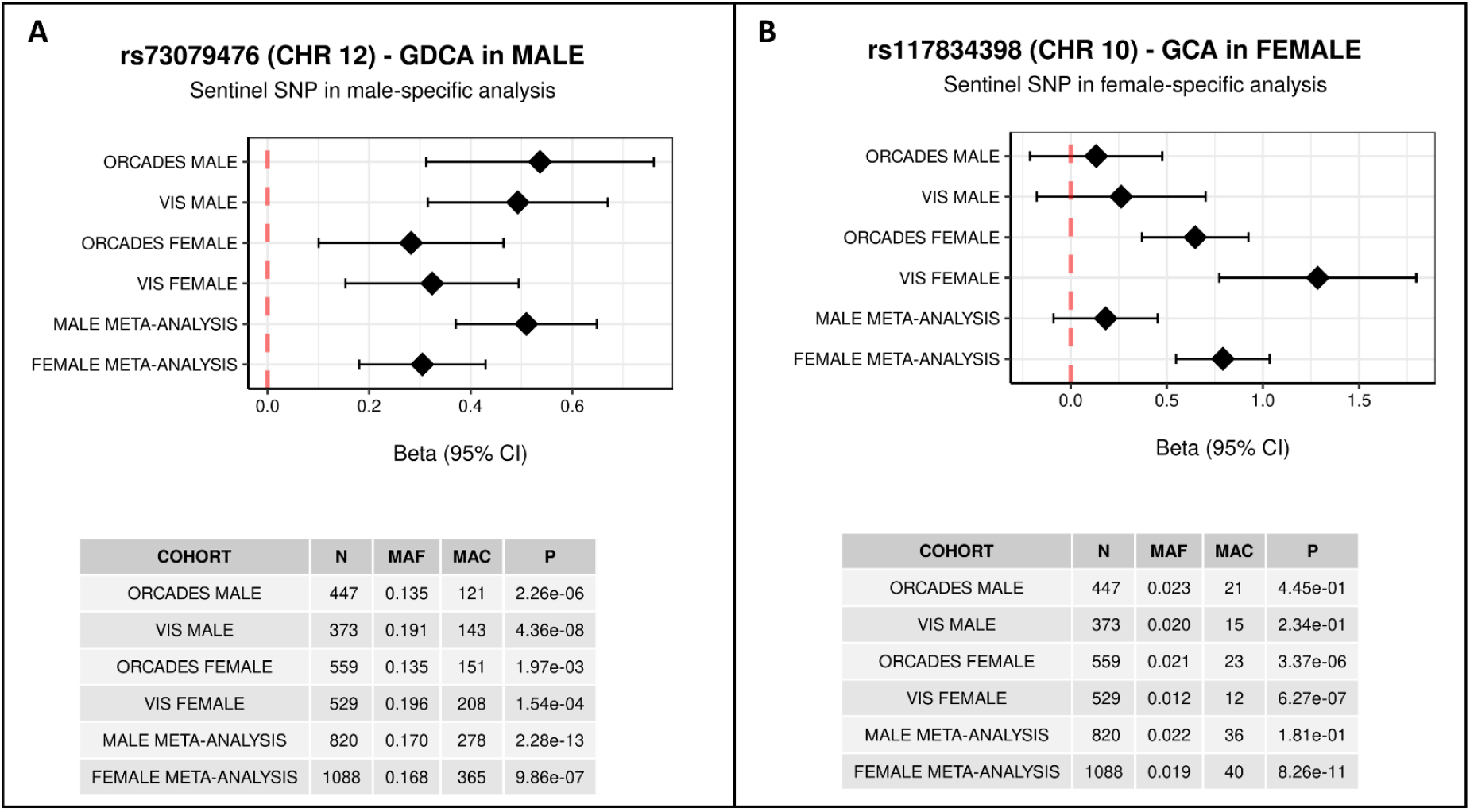
Sex-specific associations. The effect of rs73079476 on chromosome 12 on GDCA bile acid is almost as twice strong in males compared to the effect in females (Panel A). The effect of rs117834398 on GCA bile acid is stronger in females than in males (Panel B). N – sample size, MAF – minor allele frequency, MAC – minor allele count, CI – confidence interval.

### Link with complex traits and diseases

Next, we assessed whether variants associated with BA levels have been previously associated with any other biochemical traits and diseases. Using Phenoscanner^16,17^ we found that rs4149056, sentinel SNP in *SLCO1B1* locus, and its proxies (r^2^ > 0.8), were also associated with concentration of bilirubin, non-bile acid metabolites, mean corpuscular haemoglobin, sex hormone binding globulin and estrone conjugates, and various responses to drugs (i.e., statin-induced myopathy, LDL-cholesterol response to simvastatin and methotrexate clearance in acute lymphoblastic leukaemia) (Supplementary Table 4). To obtain deeper insight into the causal relationship between BAs and diseases, we conducted bi-directional Mendelian Randomisation (MR) analysis. Using the sentinel SNPs associated with GLCA, GDCA and GCA (Table 1) as instrumental variables we tested whether genetically increased levels of BA influence levels or risk for 548 biochemical traits and diseases available in the IEU Open GWAS database^18^ (Supplementary Table 5). Levels of GLCA and GDCA were significantly (p-value < 0.05/(548×3) = 3.04×10^−5^) associated with different biochemical measurements, such as levels of sex hormone-binding globulin, testosterone, triglycerides, vitamin D, alanine transaminase and galectin-3; with blood traits, such as mean corpuscular haemoglobin and mean corpuscular volume; and with diseases and their risk factors, such as daytime dozing and stroke (Supplementary Table 6). These MR tests were performed using the Wald ratio test utilising only a single instrument, thus the results of causal relationship between BAs and traits/diseases should be interpreted with caution. Yet our results suggest a possible overlap in genetic regulation, involving the *SLCO1B1* locus. Next, to assess whether complex traits and disease could have an effect on bile acid levels, we performed reverse MR using 548 traits/diseases as exposure and bile acids as outcomes. We found no significant associations, suggesting that none of the tested diseases or complex traits have an effect on BA levels (Supplementary Table 7).

### Exome-wide rare variant analysis of bile acids

To assess the contribution of low frequency and rare variants to the bile acid genetic architecture, we performed exome-wide gene-based tests across 18 bile acid traits in the ORCADES cohort (N = 1006) by testing the aggregated effect of rare (MAF <5%) predicted loss-of-function (pLoF) and non-synonymous missense variants. We identified significant association (p-value <1.79 × 10^−7^) of rare variants from 3 genes with 2 bile acid traits (quantitative CA and binary THDCA). For these associations, a significant p-value was reported by at least 2 of the 4 aggregation tests used. Rare variants significantly associated with quantitative bile acid trait CA are located in the *OR1G1* gene, while those associated with binary bile acid trait THDCA are located in *SART1* and *SORCS2* genes (Table 2, Supplementary Table 8). We further identified significant association of rare variants from *EPS8L1* gene with quantitative bile acid trait DCA and from *EEF2K* with binary bile acid trait THDCA (Supplementary Table 8). However, a significant p-value was reported by only one of the 4 aggregation tests used. Due to the lack of replication across aggregations tests, we considered these associations as not robust.

**Table 2.**
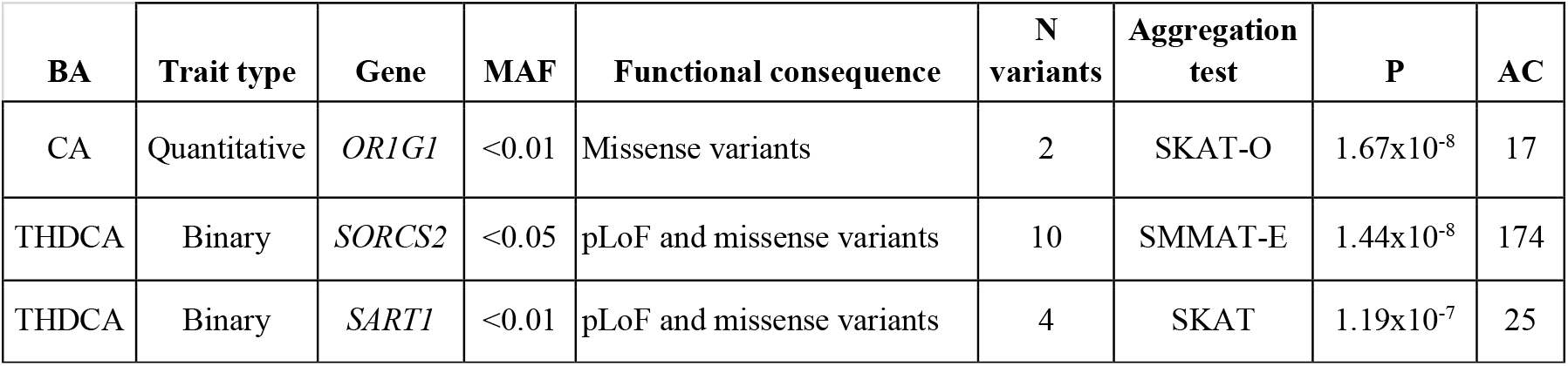
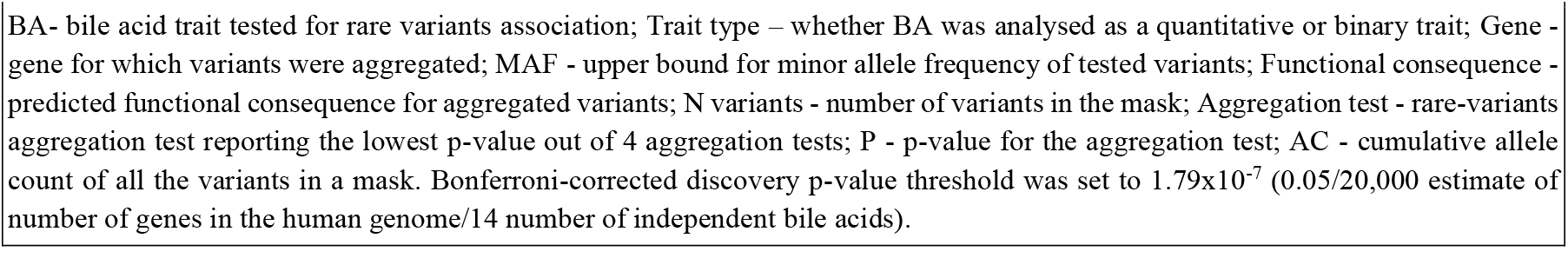
Gene-based aggregation analysis results for bile acid traits in ORCADES cohort.

## Discussion

Bile acids (BAs) are synthesised from cholesterol in the liver and then secreted into the small intestine to emulsify and promote absorption of lipid-soluble nutrients. BAs also act as hormone-like signalling molecules and have been linked to regulation of lipid and glucose metabolism, immunity, vitamin absorption, hepatobiliary diseases, inflammatory bowel disease and cancer. Despite the crucial role of BAs on whole-body physiology, their genetic architecture has not been extensively investigated in a large sample from the general population. In this study, we performed both pooled and sex-stratified genome-wide association meta-analysis of plasma levels of 18 bile acid compounds, including both primary and secondary forms, in 4923 European individuals.

We identified two secondary bile acids (GDCA and GLCA) significantly associated with a locus encompassing the *SLCO1B1* gene. The encoded protein, OATP1B1 (organic anion transporting polypeptide 1B1), is a well-known human hepatocyte transporter mediating the uptake of various endogenous compounds such as bile salts, bilirubin glucuronides, thyroid hormones and steroid hormone metabolites, and also clinically frequently used drugs like statins, HIV protease inhibitors, and the anti-cancer agents irinotecan or methotrexate^19–23^. The sentinel SNP of the *SLCO1B1* locus, rs4149056, is a missense variant (p.Val174Ala) which has been linked by previous GWA studies to blood concentration of several metabolites, including vitamin D^24^, triglycerides^25^ and bilirubin^26^, a compound resulting from the breakdown of haem catabolism and excreted as a major component of bile. This same variant has also been associated with levels of sex hormone-binding globulin and testosterone^27^. The knock-out of the gene in mice results in abnormal liver physiology and abnormal xenobiotic pharmacokinetic phenotypes (Open Targets^28^). A rare variant from the *PRKG1* locus was significantly associated with levels of glycocholic acid (GCA). *PRKG1* encodes a Protein Kinase CGMP-Dependent 1, a protein involved in signal transduction and a key mediator of the nitric oxide/cGMP. The sentinel variant in the region, rs146800892, only passes the MAF threshold (MAF > 0.01) in the CROATIA-Vis cohort, which is therefore the only cohort contributing to this association. Due to its demographic history and geographic position, CROATIA-Vis is a genetic isolate^29^ so it is possible that this variant has increased in frequency compared to a general population^30^.The mechanism of how the variation within this gene could relate to bile acid levels is unclear and would need to be further investigated.

In the sex-stratified GWAS meta-analysis, we observed sex-specific associations for the two identified loci. Levels of glycodeoxycholic acid (GDCA) are more strongly associated with the variant in *SLCO1B1* in men than in women, while female levels of GCA are more strongly affected by the variant in *PRKG1* than male levels. Later, our Mendelian randomization analysis did not provide evidence that testosterone, oestradiol, sex hormone-binding globulin or other sex-related traits have causal effects on plasma BA levels. While this could be due to a lack of statistical power of our BA meta-analysis, we currently have no evidence to suggest an effect of sex-related hormones on BA levels mediated by genetics. We also detected associations with variants from the same gene, *PRKG1*, in the main, non-stratified analysis. However, the two associations (sex-specific and pooled) appear to be independent (LD r^2^ <0.001). While the association from the pooled analysis might be either false positive or population-specific, the independent association from the sex-stratified analysis replicates well between two analysed cohorts (CROATIA-Vis and ORCADES).

After assaying common variants through GWAS, we performed exome-wide gene-based association tests in a subset of our samples (N = 1006), to investigate the genetic contribution of rare and low frequency (MAF <5%) coding variants (pLoF and missense) to bile acid levels. Overall, we identified associations with rare variants from 3 genes, *OR1G1, SART1* and *SORCS2. OR1G1* is an olfactory receptor gene, whose coded protein receptor interacts with odorant molecules in the nose to initiate a neuronal response triggering the perception of smell^31,32^. In addition to the nasal level, the olfactory receptor coded by *OR1G1* is expressed also by enterochromaffin cells, specialised enteroendocrine cells of the gastrointestinal tract. Braun *et al*^33^. determined that certain olfactory cues from spices and odorants, such as thymol, present in the luminal environment of the gut may stimulate serotonin release via olfactory receptors present in enterochromaffin cells. Between 90% and 95% of total body serotonin is in fact synthesised by enterochromaffin cells^34^: serotonin controls gut motility and secretion and is implicated in pathologic conditions such as vomiting, diarrhoea, and irritable bowel syndrome^33^. In mice, gut serotonin was shown to stimulate bile acid synthesis and secretion by the liver and gallbladder. Thus, release of serotonin in response to odorant cues increases bile acid turnover^35^. The hypoxia-associated factor (HAF), encoded by *SART1* gene and also known as SART1(800), is involved in proliferation and hypoxia-related signalling. The protein encoded by *SORCS2* is a receptor for the precursor of nerve growth factor, up-regulation of which has been reported for several liver pathologies, such as hepatotoxin-induced fibrosis^36^, ischemia-reperfusion injury^37^, oxidative injury^38^, cholestatic injury^39^ and hepatocellular carcinoma^36,40,41^. However, due to unavailability of exome sequencing data in other cohorts these associations were not replicated.

Recently, Chen *et al*.^42^ have performed an association analysis on plasma and faecal levels of bile acids in 297 obese individuals. Their study revealed 27 associated loci, including genes involved in transport of GDP-fucose and zinc/manganese and zinc-finger-protein-related genes, mostly associated with bile acid levels in stool. In our study we analysed blood plasma in a much larger sample from a general population and discovered only two associated loci. Neither of genes identified in our study were reported in Chen *et al*, suggesting that genetic regulation of bile acids between stool and blood plasma or between obese and general populations might differ significantly.

We acknowledge several limitations in the present study. We found only a small percentage of BA variability to be affected by genetics, suggesting that a larger sample size is required to further describe BA genetic architecture. BAs are known to be largely influenced by environmental factors, such as sex and gut microbiota. Female sex and oestrogens are considered relevant regulators of BA production and composition^43,44^. In pregnant women, high levels of circulating oestrogen are associated with development of cholestasis, characterised by increased serum bile acids, likely via oestrogen reducing the expression of BA receptor and transport proteins^45^. Similarly, age-related differences in hormone levels influence the differential production of BAs in women^46^. The relevant impact of sex on plasma BA levels was confirmed by the sex-stratified analysis, where the two significantly associated loci showed to be sex-specific. Similarly, species-composition of gut microbiota has a great impact on BAs levels, especially for secondary BAs that are a direct result of microbiome activity. A recent study describing the effect of gut microbiota on the human plasma metabolome reported that both primary BA cholic acid (CA) and secondary BA deoxycholic acid (DCA) show a high percentage of variance explained by the microbiota (R^2^ = 30% and 36%, respectively), indicating a strong impact on BAs of the variation in microbiota composition^47^. It is important to interpret our findings in the context of the tissue in which BA levels were measured, blood plasma. Bile acids are synthesised in the liver and secreted into the intestine, to be then reabsorbed into portal circulation and returned to the liver: plasma BA levels thus reflect the amount of BAs escaping extraction from the portal blood. Therefore, levels of BAs in plasma are likely to be influenced by genes other than those encoding the particular anabolic and catabolic enzymes, including those involved in hepatic function and dysfunction. In line with this, the major genetic contributor to blood BA levels in our study are variants from the *SLCO1B1* gene, encoding the hepatocyte transporter OATP1B1 and important for flux of bile salts, bilirubin glucuronides and various hormone metabolites, rather than genes encoding key enzymes of primary BA synthesis, such as *CYP7A1* and *CYP7B1*^48^. Similarly, some of the genes with rare variants associations have been linked to liver diseases, such as liver cancer^49^, and intrahepatic cholestasis of pregnancy^50^.

In conclusion, we explored the genetic architecture of plasma bile acid levels, including both common and rare variants. By performing GWAS meta-analysis (N = 4923), we identified 2 significantly associated loci, mapping to the *SLCO1B1* and *PRKG1* genes. In the sex-specific GWAS meta-analysis we observed that variants in these genes have different impact on bile acid levels in men and women. To assess relationships between genetically increased levels of bile acids and risk for diseases we performed Mendelian randomisation, but did not find any bile acids affecting disease risk, nor the reverse, which however might be affected by the lack of statistical power. Using the gene-based aggregated tests and whole exome sequencing, we further identified rare pLoF and missense variants in 3 genes associated with BAs, *OR1G1, SART1* and *SORCS2*, some of which are known to be involved in liver disease. Additional studies with larger sample sizes and of more diverse ancestry will be necessary to validate our findings, further unravel the genetic architecture of bile acid levels, and to understand their relationship with human diseases and complex traits.

## Materials and methods

### Phenotypic data

#### Bile acids quantification

Bile acid (BA) analysis was performed from plasma or serum (MICROS cohort) samples by liquid chromatography-tandem mass spectrometry (LC-MS/MS) as previously described^51^. The HPLC equipment consisted of a 1200 series binary pump (G1312B), a 1200 series isocratic pump (G1310A) and a degasser (G1379B) (Agilent, Waldbronn, Germany) connected to an HTC Pal autosampler (CTC Analytics, Zwingen, CH). A hybrid triple quadrupole linear ion trap mass spectrometer API 4000 Q-Trap equipped with a Turbo V source ion spray operating in negative ESI mode was used for detection (Applied Biosystems, Darmstadt, Germany). High purity nitrogen was produced by a nitrogen generator NGM 22-LC/MS (cmc Instruments, Eschborn, Germany). Gradient chromatographic separation of BAs was performed on a 50 mm × 2.1 mm (i.d.) Macherey-Nagel NUCLEODUR C18 Gravity HPLC column, packed with 1.8 μm particles equipped with a 0.5 μm pre-filter (Upchurch Scientific, Oak Harbor, WA, USA). The injection volume was 5 μL and the column oven temperature was set to 50 °C. Mobile phase A was methanol/water (1/1, v/v), mobile phase B was 100% methanol, both containing 0.1% ammonium hydroxide (25%) and 10 mmol/L ammonium acetate (pH 9). A gradient elution was performed with 100% A for 0.5 min, a linear increase to 50% A until 4.5 min, followed by 0% A from 4.6 until 5.5 min and re-equilibration from 5.6 to 6.5 min with 100% A. The flow rate was set to 500 μL/min. To minimize contamination of the mass spectrometer, the column flow was directed only from 1.0 to 5.0 min into the mass spectrometer using a diverter valve. Otherwise, methanol with a flowrate of 250 μL/min was delivered into the mass spectrometer. The turbo ion spray source was operated in the negative ion mode using the following settings: Ion spray voltage = −4500 V, ion source heater temperature = 450 °C, source gas 1 = 40 psi, source gas 2 = 35 psi and curtain gas setting = 20 psi. Analytes were monitored in the multiple reaction monitoring (MRM). Quadrupoles Q1 and Q3 were working at unit resolution. Calibration was achieved by the addition of BAs to EDTA-plasma/serum. A combined BA standard solution containing the indicated amounts (0.5 - 70.5 μmol/L) was placed in a 1.5 ml tube and excess solvent was evaporated under reduced pressure before adding EDTA-plasma/serum. Calibration curves were calculated by linear regression without weighting. Data analysis was performed with Analyst Software 1.4.2. (Applied Biosystems, Darmstadt, Germany). The data were exported to Excel spreadsheets and further processed by self-programmed Excel macros which sort the results, calculate the analyte/internal standard peak area ratios, generate calibration lines and calculate sample concentrations. For the calculation we selected the internal standard with analogous fragmentation and closest retention time to the respective BA species.

#### Pre-processing of bile acid traits

Prior to genetic analysis, bile acid traits were grouped into three groups based on the percentage of samples with below the limit of detection (<LOD) measurements: high <LOD group (> ∼30% of all samples below LOD) and low <LOD group (< ∼7% of all samples below LOD) (Supplementary Table 1). Accordingly, different phenotypic pre-processing and different analysis strategies were applied to the groups. Bile acids within a high <LOD were considered as binary traits: individuals were categorised based on whether their bile acid levels were effectively measured (category 1) or were below the LOD (category 0). Bile acid traits belonging to this group were THDCA, TUDCA, TCA and GHDCA. All other bile acids were considered as quantitative traits and were log_10_-transformed. However, to increase the sample size, in addition to a complete-case analysis (considering as missing all samples with <LOD), we also imputed <LOD measurements. For each bile acid, imputation of <LOD measurements was performed by fitting a truncated normal distribution, with mean and standard deviation of the effectively measured raw data, truncated (as an upper bound) to the lowest measured value for the given bile acid. To do so, we used the “rtnorm” function from the MCMCglmm R package^52^. After imputation, measurements were log_10_-transformed.

### Genome-wide association analysis

Genome-wide association studies (GWAS) were performed in 5 cohorts of European descent, CROATIA-Vis (N=971), ORCADES (Orkney Complex Disease Study) (N=1019), NSPHS (Northern Sweden Population Health Study) (N=718), MICROS (Micro-Isolates in South Tyrol) (N=1336) and ERF (Erasmus Rucphen Family Study) (N=879), for a combined sample size of 4923. Specific sample size for each bile acid molecule, in both meta-analysis and single cohort GWAS, can be found in Supplementary Table 10. Bile acid traits were adjusted for age, sex, batch, population structure/cryptic relatedness by including population principal components or applying linear mixed models and using a kinship matrix estimated from genotyped data. Within each cohort, residuals of covariate and population structure/relatedness correction were tested for association with Haplotype Reference Consortium (HRC)^53^ imputed SNP dosages or SNP genotypes from whole genome sequencing, applying an additive genetic model of association. Details of cohorts, individual-level pre-imputation QC, GWAS software and parameters specific for each cohort can be seen in Supplementary Table 11 Single-cohort summary statistics were filtered for minor allele frequency (MAF) > 0.01. The genomic control inflation factor (λ_GC_) was calculated for each bile acid trait. Cohort-level λ_GC_ overall ranged from 0.9 to 1.1 for quantitative bile acid traits, both imputed and not, suggesting little residual influence of population stratification and family structure (Supplementary Table 12). In a few cases, ERF cohort reported somewhat deflated λ_GC_ (GCDCA at 0.884 and GLCA at 0.899). On the other hand, there was considerable inflation for binary bile acid in the case of NSPHS (Supplementary Table 12), with values of λ_GC_ above 1.1, suggesting that population structure/cryptic relatedness was not fully controlled for these traits in the NSPHS cohort.

### Meta-analysis

Prior to meta-analysis, cohort-level GWAS were quality controlled using the EasyQC software package, following the protocol described in Winkler *et al*.^54^ Cohort-level results were corrected for the genomic control inflation factor, then pooled and analysed with METAL v2011-03-25 software^55^, applying the fixed-effect inverse-variance method. The mean genomic control inflation factor after the meta-analysis was 0.991 (range 0.938 – 1.009), suggesting that the confounding effects of the family structure were correctly accounted for (Supplementary Table 12). The standard genome-wide significance threshold was Bonferroni corrected for the number of independent bile acid traits, calculated as 14 (5×10^−8^/14 = 3.57×10^−9^). The number of independent bile acid traits was estimated as the sum of the number of binary traits (4) and the number of principal components that jointly explained 99% of the total variance of log_10_-transformed quantitative traits in each cohort (10) (Supplementary Table 13).

### Sex-stratified GWAS meta-analysis

To identify possible differences in the genetic contribution to bile acid variability between men and women, we performed sex-specific GWAS of the 14 quantitative bile acid traits for ORCADES and CROATIA-Vis cohorts. Given that for the sex-stratified GWAS we implicitly halve our sample size, we performed these analyses only on the imputed bile acid traits. The same analysis steps and procedures already described for the full meta-analysis were applied. Bile acid traits were adjusted for age, sex and batch as fixed effects, and relatedness (estimated as the kinship matrix calculated from genotyped data) as a random effect in a linear mixed model, calculated using the ‘polygenic’ function from the GenABEL R package^56^. Residuals of covariate and relatedness correction were tested for association with HRC-imputed^53^ SNP dosages using the RegScan v0.5 software^57^, applying an additive genetic model of association. Prior to meta-analysis, SNPs having a difference in allele frequency between the two cohorts higher than ±0.3 or a minor allele count (MAC) lower or equal to 6 were filtered out. Cohort-level GWAS were corrected for genomic control inflation factor and then meta-analysed (N =820 for male and N =1088 for female individuals) using METAL v2011-03-25 software^55^, applying the fixed-effect inverse-variance method. The mean λ_GC_ was 0.993 (range 0.978– 1.011) for male-specific meta-analysis and 0.996 (range 0.984–1.003) for female-specific meta-analysis. The Bonferroni-corrected significance threshold applied is 5 × 10^−9^.

### Phenoscanner and Mendelian Randomisation

To assess link between bile acids and diseases we explored the overlap of SNPs associated with BAs with complex human traits by using PhenoScanner v1.1 database^16,17^, taking into account significant genetic association (*p* < 5 × 10^−9^) at the same or strongly (LD *r*2 > 0.8) linked SNPs in populations of European ancestry. We then performed bi-directional Mendelian Randomisation (MR) to investigate the effect of 548 complex traits and diseases available in the IEU Open GWAS database^18^ (manually curated list of studies from identifiers ebi-a, ieu-a, ieu-b and ukb-a; the complete list reported in the Supplementary Table 5) on BA levels, and vice-versa. The set of genome-wide significant, LD clumped SNPs used as instruments for complex traits/diseases was extracted from the selected studies by using the “<u>extract_instruments</u>“ function from the TwoSampleMR 0.5.6 R package^58^. Similarly, sentinel SNPs from BAs meta-analysis (Supplementary Table 2) were selected as instruments. MR tests were performed by using fixed effects inverse variance-weighted (IVW) in case of multiple instruments or Wald Ratio method in case of a single instrument, as implemented in the TwoSampleMR 0.5.6 R package^58^. Multiple testing correction was controlled for using either the Bonferroni correction or false discovery rate (FDR).

### Whole-exome sequencing data

#### Exome sequencing

The “Goldilocks” exome sequence data for ORCADES cohort was prepared at the Regeneron Genetics Center, following the protocol detailed in Van Hout *et al*.^59^ for the UK Biobank whole-exome sequencing project. In summary, sequencing was performed using S2 flow cells on the Illumina NovaSeq 6000 platform with multiplexed samples. DNAnexus platform^60^ was used for processing raw sequencing data. The files were converted to FASTQ format and aligned using the BWA-mem^61^ to GRCh38 genome reference. The Picard tool^62^ was used for identifying and flagging duplicated reads, followed by calling the genotypes for each individual sample using the WeCall variant caller^63^. During quality control, 33 samples genetically identified as duplicates, 3 samples showing disagreement between genetically determined and reported sex, 4 samples with high rates of heterozygosity or contamination, 2 samples having low sequence coverage (less than 80% of targeted bases achieving 20X coverage) and 1 being discordant with genotyping chip were excluded. Finally, the “Goldilocks” dataset was generated by (i) filtering out genotypes with read depth lower than 7 reads, (ii) keeping variants having at least one heterozygous variant genotype with allele balance ratio greater than or equal to 15% (AB ≥ 0.15) or at least one homozygous variant genotype, and (iii) filtering out variants with more than 10% of missingness and HWE p<10^−6^. Overall, a total of 2,090 ORCADES (820 male and 1,270 female) participants passed all exome sequence and genotype quality control thresholds. A pVCF file containing all samples passing quality control was then created using the GLnexus joint genotyping tool^64^.

#### Variant annotation

Exome sequencing variants were annotated as described in Van Hout, *et al*.^59^ Briefly, they were annotated with the most severe consequence across all protein-coding transcripts using SnpEff^65^. Gene regions were defined based on Ensembl release 85^66^. Predicted loss-of function (pLoF) variants were defined as variants annotated as start lost, stop gained/lost, splice donor/acceptor and frameshift. The deleteriousness of missense variants was based on dbNSFP 3.2^67,68^ and assessed using the following algorithms: (1) SIFT^69^: “D” (Damaging), (2) Polyphen2_HDIV: “D” (Damaging) or “P” (Possibly damaging), (3) Polyphen2_HVAR^70^: “D” (Damaging) or “P” (Possibly damaging), (4) LRT^71^: “D” (Deleterious) and (5) MutationTaster^72^: “A” (Disease causing automatic) or “D” (Disease causing). If not predicted as deleterious by any of the algorithms the missense variants were considered “likely benign”, “possibly deleterious” if predicted as deleterious by at least one of the algorithms and “likely deleterious” if predicted as deleterious by all five algorithms.

### Exome-wide gene-based aggregation analysis of rare variants

#### Generation of gene masks

For each gene, the variants were grouped into four categories (masks), based on severity of their functional consequence. The first mask (mask 1) included only pLoF variants. Masks 3 and 4 included both pLoF and variants predicted to be deleterious, by 5/5 algorithms (mask 3) or by at least one algorithm (mask 4). The most permissive mask (mask 2) included pLoF and all missense variants. These masks were then further split by the frequencies of the minor allele (MAF ≤ 5%, e.g. mask1_maf5; and MAF ≤ 1%, e.g. mask1_maf1), resulting in up to 8 burden tests for each gene (Supplementary Table 9).

#### ORCADES gene-based aggregation analysis

We performed variant Set Mixed Model Association Tests (SMMAT)^73^ on the 18 bile acid traits from ORCADES cohort, quantified and pre-processed as previously described, fitting a GLMM adjusting for age, sex, batch, and familial or cryptic relatedness by kinship matrix. The kinship matrix was estimated from the genotyped data using the ‘ibs’ function from GenABEL R package^56^. The SMMAT framework includes 4 variant aggregate tests: burden test, sequence kernel association test (SKAT), SKAT-O and SMMAT-E, a hybrid test combining the burden test and SKAT. The 4 variant aggregate tests were performed on 8 different pools of genetic variants, called “masks”, each one including a different set of variants based on both MAF and predicted consequence of variants (e.g., loss of function and missense) (Supplementary Table 9), as described above. Discovery significance threshold was Bonferroni corrected for the rough estimate of the number of genes in the human genome, 20,000, and the number of independent bile acid traits, 14, calculated as previously described (0.05/20000/14 = 1.79×10^−7^). A gene association was considered significant if it passed the above reported Bonferroni corrected significance threshold in at least two of the 4 performed variant aggregate tests and if the cumulative allele count of the variants included in the gene was equal or higher than 10.

## Data Availability

The full summary statistics from GWAS meta-analysis of bile acids will be uploaded to the University of Edinburgh Datashare repository and to GWAS catalog upon manuscript acceptance. There is neither Research Ethics Committee approval, nor consent from individual participants, to permit open release of the individual level research data underlying this study. The datasets analysed during the current study are therefore not publicly available. Instead, the research data and/or DNA samples for the ORCADES study are available from on reasonable request, following approval by the QTL Data Access Committee and in line with the consent given by participants. Each approved project is subject to a data or materials transfer agreement (D/MTA) or commercial contract. The summary statistics for complex traits and diseases (full list reported in Supplementary Table 5) are available in the IEU Open GWAS database https://gwas.mrcieu.ac.uk/.

## Code availability

We used publicly available software tools for all analyses. These software tools are listed in the main text and in the Methods.

## Acknowledgments

The Orkney Complex Disease Study (ORCADES) was supported by the Chief Scientist Office of the Scottish Government (CZB/4/276, CZB/4/710), a Royal Society URF to J.F.W., the MRC Human Genetics Unit quinquennial programme “QTL in Health and Disease”, Arthritis Research UK and the European Union framework program 6 EUROSPAN project (contract no. LSHG-CT-2006-018947). DNA extractions were performed at the Edinburgh Clinical Research Facility, University of Edinburgh. We would like to acknowledge the invaluable contributions of the research nurses in Orkney, the administrative team in Edinburgh and the people of Orkney. For the purpose of open access, the author has applied a Creative Commons Attribution (CC BY) licence to any Author Accepted Manuscript version arising from this submission. The CROATIA-VIS study in the Croatian island of Vis was supported through the grants from the Medical Research Council UK and Ministry of Science, Education and Sport of the Republic of Croatia (number 108-1080315-0302). The authors collectively thank a large number of individuals for their individual help in organising, planning and carrying out the field work related to the project and data management: Professor Pavao Rudan and the staff of the Institute for Anthropological Research in Zagreb, Croatia (organisation of the field work, anthropometric and physiological measurements, and DNA extraction); Professor Ariana Vorko-Jovic and the staff and medical students of the Andrija Stampar School of Public Health of the Faculty of Medicine, University of Zagreb, Croatia (questionnaires, genealogical reconstruction and data entry); Dr Branka Salzer from the biochemistry lab “Salzer”, Croatia (measurements of biochemical traits); local general practitioners and nurses (recruitment and communication with the study population); and the employees of several other Croatian institutions who participated in the field work, including but not limited to the University of Rijeka and Split, Croatia; Croatian Institute of Public Health; Institutes of Public Health in Split and Dubrovnik, Croatia. SNP Genotyping of the Vis samples was carried out by the Genetics Core Laboratory at the Wellcome Trust Clinical Research Facility, WGH, Edinburgh.The MICROS (Micro-Isolates in South Tyrol) study is part of the genomic health care program ‘GenNova’ and was carried out in three villages of the Val Venosta on the populations of Stelvio, Vallelunga and Martello. We thank the primary care practitioners Raffaela Stocker, Stefan Waldner, Toni Pizzecco, Josef Plangger, Ugo Marcadent and the personnel of the Hospital of Silandro (Department of Laboratory Medicine) for their participation and collaboration in the research project. In South Tyrol, the study was supported by the Ministry of Health and Department of Educational Assistance, University and Research of the Autonomous Province of Bolzano and the South Tyrolean Sparkasse Foundation. The Northern Swedish Population Health Study (NSPHS) was funded by the Swedish Medical Research Council (project number K2007-66X-20270-01-3), and the Foundation for Strategic Research (SSF). The NSPHS as part of EUROSPAN (European Special Populations Research Network) was also supported by European Commission FP6 STRP grant number 01947 (LSHGCT-2006-01947). This work was also supported by the Swedish Society for Medical Research (ÅJ). The authors are grateful for the contribution of district nurse Svea Hennix for data collection and Inger Jonasson for logistics and coordination of the health survey. Finally, the authors thank all the community participants for their interest and willingness to contribute to the study. The Erasmus Rucphen Family (ERF) study was supported by grants from The Netherlands Organisation for Scientific Research (NWO), Erasmus MC, the Centre for Medical Systems Biology (CMSB) and the European Community’s Seventh Framework Programme (FP7/2007-2013), ENGAGE Consortium, grant agreement HEALTH-F4-2007-201413. We are grateful to all general practitioners for their contributions, Cornelia van Duijn and Ben Oostra for setting-up the ERF study, Petra Veraart for sorting out the genealogy records, Jeannette Vergeer and Peter Snijders for help in retrieving the materials needed to analyse data. We acknowledge support from the European Union’s Horizon 2020 research and innovation programme IMforFUTURE (A.L.: H2020-MSCA-ITN/721815); the RCUK Innovation Fellowship from the National Productivity Investment Fund (L.K.: MR/R026408/1) and the MRC Human Genetics Unit programme grant, ‘QTL in Health and Disease’ (J.F.W. and C.H.: MC_UU_00007/10).

## Ethics

All studies were approved by local research ethics committees and all participants have given written informed consent. The ORCADES study was approved by the NHS Orkney Research Ethics Committee and the North of Scotland REC. The CROATIA-Vis study was approved by the ethics committee of the medical faculty in Zagreb and the Multi-Centre Research Ethics Committee for Scotland. The Northern Swedish Population Health Study (NSPHS) was approved by the local ethics committee at the University of Uppsala (Regionala Etikprövningsnämnden, Uppsala). The MICROS study was approved by the ethical committee of the Autonomous Province of Bolzano, Italy. The ERF study was approved by the Erasmus institutional medical-ethics committee in Rotterdam, The Netherlands.

## Author contributions

A.L.: Data analysis and interpretation, visualisation, writing—original draft preparation, writing—review and editing. D.G.-S.: Data analysis, writing—review and editing. Å.J.: Data analysis. S.A.: Data analysis. G.L.: Quantification of bile acids, writing—original draft preparation. C.G.: Quantification of bile acids, writing—original draft preparation. G.T.: preparation, quality control and annotation of whole-exome sequencing data. A.R.S.: Funding, writing—review and editing. A.A.H.: Genomic and demographic data provider for MICROS cohort. P.P.: Funding, genomic and demographic data provider for MICROS cohort. C.P.: genomic and demographic data provider for MICROS cohort. H.C. Funding. O.P.: Genomic and demographic data provider for CROATIA-Vis cohort. C.H.: Funding, genomic and demographic data provider for CROATIA-Vis cohort. N.P.: supervision and data interpretation for bile acid pre-processing and imputation. M.G.: Genomic and demographic data provider for ERF cohort, writing—review and editing. U.G.: Funding, genomic and demographic data provider for NSPHS cohort. C.F.: Genomic and demographic data provider for MICROS cohort. J.F.W.: Funding, conceptualisation, genomic and demographic data provider for ORCADES cohort, supervision, data interpretation, writing—original draft preparation, writing—review and editing. L.K.: Conceptualisation, supervision, data interpretation, writing—original draft preparation, writing—review and editing.

