## Supplementary Figure for "Genome-wide association study reveals loci with sex-specific effects on plasma bile acids"

### Supplementary Figures

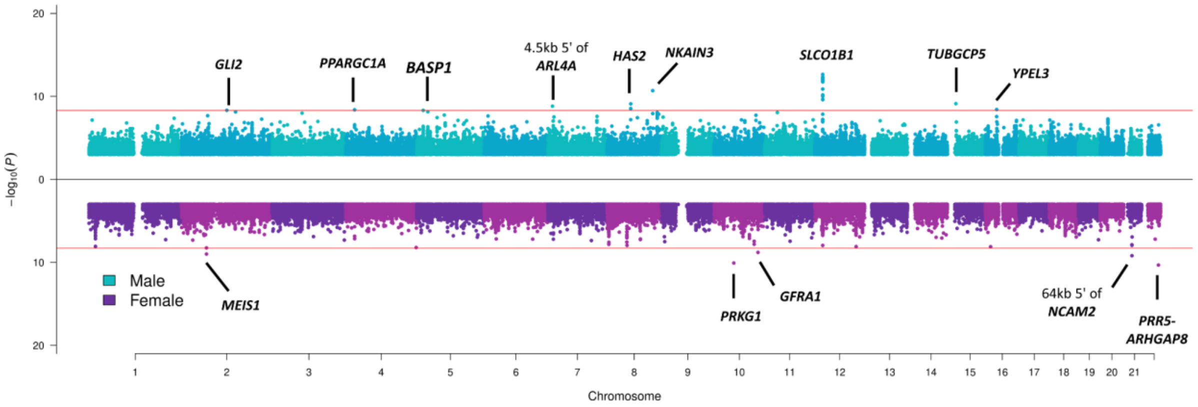


**Supplementary Figure 1. Miami plot pooling together sex-specific meta-analysis results obtained across 14 bile acid traits (quantitative traits with imputed LOD values, as described in Methods), for male at the top in blue, and for female at the bottom in purple.** The pooling was performed by selecting the lowest *p* value (y-axis) from the 14 bile acids for every genomic position (x axis). The Bonferroni-corrected genome-wide significance threshold (horizontal red lines) corresponds to $5 \times 10^{-9}$. For simplicity, SNPs with *p* value $> 1 \times 10^{-3}$ are not plotted. *P* values are derived from the two-sided Wald test with one degree of freedom.
